# Rifaximin does not increase the rate of 30-day mortality in patients with cirrhosis and daptomycin in two National US-based cohorts

**DOI:** 10.1101/2024.12.06.24318164

**Authors:** Scott Silvey, Nilang Patel, Alexander Khoruts, Jasmohan S Bajaj

## Abstract

Recently, a higher rate of resistance to daptomycin in patients exposed to rifaximin has been shown. However, since laboratory resistance patterns found *in silico* or *ex vivo* do not account for the complex pharmacokinetic, pharmacodynamic, microbial, and host-related factors in patients, the clinical impact of rifaximin on resistance to daptomycin needs clarification. Although rifaximin is commonly used for irritable bowel syndrome and traveler’s diarrhea, the main reason for long-term use is hepatic encephalopathy (HE) in cirrhosis. Here we show that 30-day outcomes (transplant or mortality) in two large US-based cohorts (Veterans Affairs Corporate Data Warehouse and TriNetX database) in daptomycin users on rifaximin were not different compared to those without daptomycin. with or without pre-existing rifaximin use.

---

Recently, a higher rate of resistance to daptomycin in patients exposed to rifaximin has been shown^1^. However, since laboratory resistance patterns found *in silico* or *ex vivo* do not account for the complex pharmacokinetic, pharmacodynamic, microbial, and host-related factors in patients, the clinical impact of rifaximin on resistance to daptomycin needs clarification^2^.

Although rifaximin is commonly used for irritable bowel syndrome and traveler’s diarrhea, the main reason for long-term use is hepatic encephalopathy (HE) in cirrhosis. These cirrhosis patients are also especially prone to suffer consequences of antibiotic resistant gram-positive and gram-negative infections^3^, which makes the study of the effectiveness of daptomycin crucial^4^. Our aim was to determine 30-day outcomes in two large US-based cohorts in daptomycin users with or without pre-existing rifaximin use. The two cohorts were the Veterans Affairs Corporate Data Warehouse (VA-CDW) and TriNetX database (Figure 1). Data were extracted from 2010-2019, since 2010 was when rifaximin was FDA approved for HE in the US, till 2019 to avoid COVID-19 related confounding. In the VA-CDW we identified cirrhosis patients who received daptomycin between 2010-2019. Rifaximin usage was defined as ≥1 prescription up to 90 days pre-index daptomycin initiation date. We collected information on demographics, medications, cirrhosis history and severity (MELD-Na, HE and ascites details, prior spontaneous bacterial peritonitis (SBP), among others], and hospital course (Supplement). The primary outcome was 30-day all-cause mortality. The above analysis was replicated within TriNetX – a national database of insured patients sourced from health care organizations (HCOs) participating in the TriNetX Research Network (trinetx.com). These HCOs are typically large non-VA academic medical institutions with both inpatient and outpatient facilities. TriNetX contains information regarding patient demographics, diagnoses and procedures through ICD or CPT codes, laboratory values, prescription data, transplant, and death records. Because the LT rate is higher in the non-VA population, we assessed three outcomes– composite 30-day mortality or liver transplant (LT), 30-day transplant-free mortality, and 30-day LT alone. Inclusion criteria were similar to VA-CDW. Detailed statistical analyses are in the supplement.

**Figure 1:**
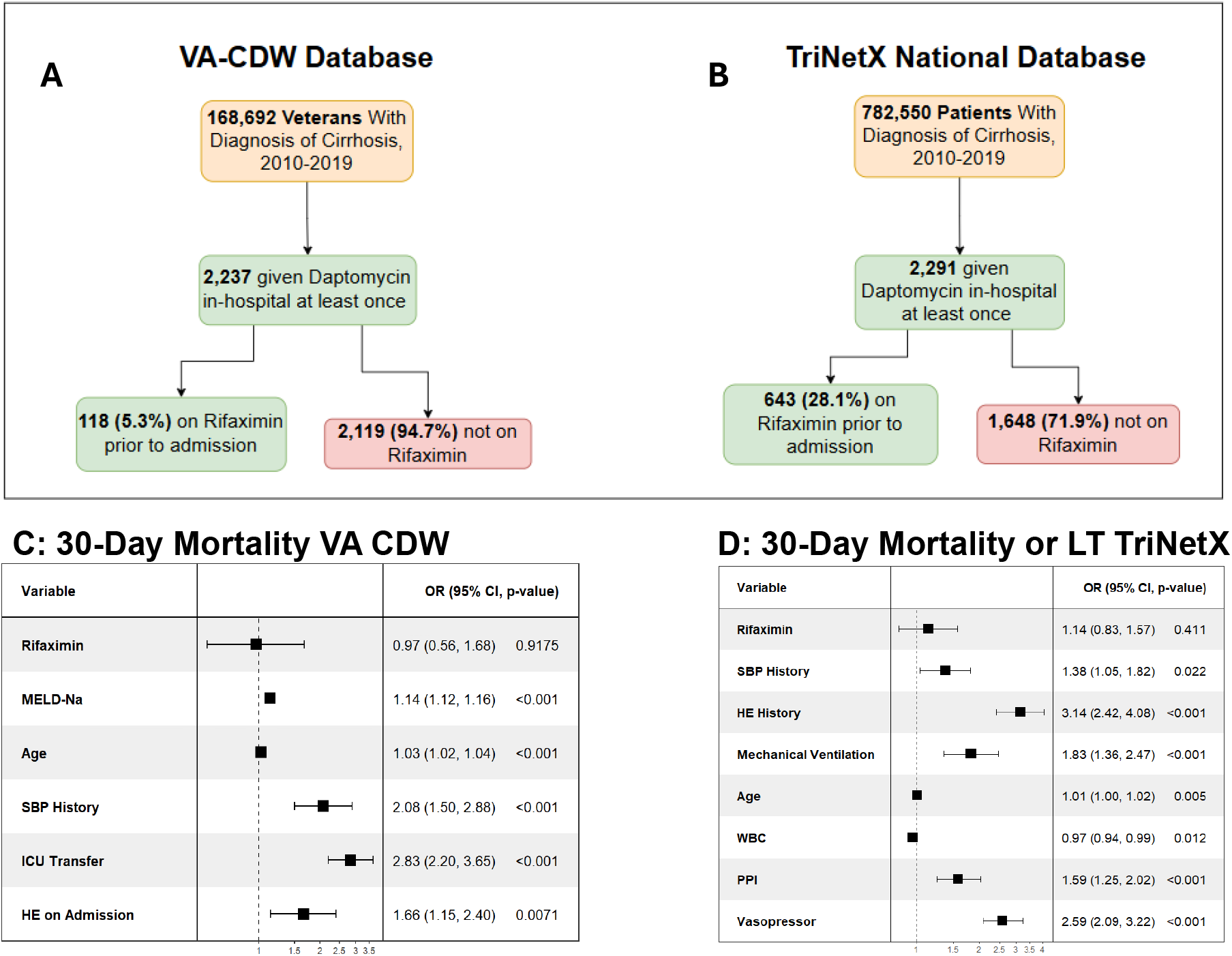
Forest Plots + Flowchart Figure legend: A: Flowchart of patients in Veterans Affairs Corporate Data Warehouse, B: Flowchart of patients in TriNetX, C: Forest plot of logistic regression in VA CDW, D: Forest plot of logistic regression in TriNetX. WBC: white blood cell count, PPI: proton pump inhibitor use, ICU: intensive care unit, HE: hepatic encephalopathy, BP: spontaneous bacterial peritonitis.

In VA-CDW we found 2237 patients that were given Daptomycin, of which 118 (5.3%) were on rifaximin prior to admission. Among these, 511 (22.8%) died within 30-days post-Daptomycin and 59 (2.6%) received LT. Cohort characteristics were compared (Table S1) - patients on rifaximin were slightly younger and were more likely to be on proton pump inhibitors (PPI), diuretics, lactulose, or beta-blockers at the time of admission. These patients also had significantly higher MELD-Na scores and were more likely to have a history of spontaneous bacterial peritonitis (SBP), variceal bleed, and chronic viral hepatitis. During their hospitalization, rifaximin users were also more likely to be transferred to the ICU. On univariable analysis, rifaximin was associated with significantly higher 30-day mortality rates (OR: 2.10 [1.42-3.08], *p*<0.001). However, after adjusting for cirrhosis severity, patient history, other medication usage, and hospital course, this association was no longer statistically significant (adjusted OR: 0.97 [0.52-1.55], *p*=0.918). A full visualization of all variables are in Figure 1C.

In TriNetX we identified 2,291 patients who were given daptomycin, of which 643 (28.1%) were on Rifaximin. Among these patients, 753 (32.9%) died or received LT, 422 (18.4%) died in 30-days post-Daptomycin without transplant and 342 (14.9%) received liver transplant. Cohort characteristics were compared (Table 1) – these were like trends seen in the VA-CDW database. On univariable analysis, rifaximin was associated with significantly higher 30-day composite mortality/LT rate (OR: 3.80 [3.14-4.60], *p*<0.001), which was again strongly diminished after adjusting for cirrhosis severity and hospital course (Figure 1D, adjusted OR: 1.14 [0.83-1.57], *p*=0.411). LT-free mortality (OR: 3.96 [3.14-5.00], *p*<0.001) was also significantly higher in rifaximin patients on univariable analysis, but again the significance disappeared with adjustment (Figure S1A). Finally, LT rates (OR: 2.52 [1.99-3.19], *p*<0.001) were again higher in rifaximin patients on univariable (24.3% vs. 11.3%) but not on multivariable analysis (Figure S1B). However, this demonstrated that rifaximin was not a barrier to LT.

This data strongly demonstrate that concomitant rifaximin use in patients with cirrhosis that are subsequently prescribed daptomycin is rare and importantly does not increase the risk of 30-day mortality across two national US-based cohorts. While AMR testing is a critical aspect of streamlining therapies, care must be taken to interpret these results in a clinical context^2^.

Rifaximin has grade A evidence to prevent HE recurrence^5^. Moreover, in some studies rifaximin has even been shown to reduce infections and in national analyses has shown to improve clinical outcomes. In some cases, rifaximin has been shown to potentially even prevent outpatient and inpatient infections without a clinically significant infection or resistant infection-related signal^6^. Importantly, rifaximin in cirrhosis patients reduces the burden of mucolytic bacteria and promotes gut barrier repair, thus enhancing protection against invading pathobionts^7, 8^. Since FDA approval, there have been several analyses that have not shown clinically relevant increases in antibiotic resistance with rifaximin^9^. Furthermore, here was no signal for selective increase in gram-positive taxa including *Enterococci* in any rifaximin study from an overall microbiome or culture-based study^10^.

Controlling for several important factors that are associated with inpatient outcomes in cirrhosis, we did not demonstrate a significant difference in rifaximin utilizing patients on daptomycin compared to daptomycin alone. Veterans are more likely to be older, less diverse and with higher comorbid conditions than the populations included in TriNetX. Therefore, similar patterns among in two national cohorts that have varying, and diverse representation is reassuring. We chose mortality due to the “last resort” status of daptomycin and included LT as an important outcome. Importantly, eligibility for LT requires stabilized or controlled infections, and in TriNetX we found that rifaximin use was not a systematic barrier to LT.

In summary, we found no significant changes in 30-day mortality or LT with the addition of rifaximin in the setting of daptomycin after controlling for important variables that are associated with cirrhosis-related mortality with or without LT. Therefore, we believe that a higher burden of proof, including translation of data presented in the recent published experience into real-world outcomes, is critical considering restriction of rifaximin in cirrhosis.

## Supporting information

Supplementary methods and results

## Data Availability

All data produced in the present study are available upon reasonable request to the authors

