## Supplementary methods and results for "Rifaximin does not increase the rate of 30-day mortality in patients with cirrhosis and daptomycin in two National US-based cohorts"

**Statistical analysis:** Cohort characteristics were summarized and compared between the groups: Rifaximin vs. No-Rifaximin. Continuous variables were presented as the mean (±SD) or median (IQR), and categorical variables as counts and percentages of the total. Variables were compared between the groups using two-sample *t*-tests, Wilcoxon Rank-Sum tests, or Pearson’s Chi-Squared tests, as appropriate. We assessed the effect of Rifaximin usage pre-daptomycin administration on each outcome described previously using logistic regression models in an univariable setting and after adjusting for all cohort characteristics that were significantly different between the treatment groups. RStudio version 4.4.1 was used for all statistical analysis. All hypothesis tests were two-sided with statistical significance considered *p*<0.05.

**Supplementary methods– Data Dictionary**

**Demographics and Prior History:**

**Cirrhosis DX (Inclusion Criteria):**

Description: History of cirrhosis prior to index SBP episode.

ICD Codes / Definition: ICD-10 : I85.00, I85.01, I85.10, I85.11, K65.2, K70.11, K70.30, K70.31, K70.40, K70.41, K71.51, K71.7, K72.10, K72.11, K74.4, K74.60. K74.69, K76.6, K76.7, K76.81

ICD-9 : 456.0, 456.1, 456.20, 456.21, 571.2, 571.5, 572.2, 572.3, 572.4

**Daptomycin (inclusion criteria):**

Description: Did the patient receive daptomycin between 2010-2019?

Definition: First date when this occurs.

**Rifaximin:** Description: Did the patient receive rifaximin between 2010-2019?

Definition: First date when this occurs.

**Age:** Patient’s age at index diagnosis.

**Male Sex:** Sex (1 = male, 0 = other).

**White Race:** Race (1 = white, 0 = other)

**Hispanic Ethnicity:** Ethnicity (1 = Hispanic/Latino, 0 = other).

**Alcohol-related Cirrhosis:**

Alcohol etiology of cirrhosis (1 = yes, 0 = no).

ICD Codes: ICD-9 : 571.2, 571.1, 571.3

ICD-10 : K70.11, K70.30, K70.31, K70.40, K70.41, K70.9, K70.10

**MELD-Na:** Patient’s most recent MELD-Na value around index diagnosis, up to 6-months pre-index.

**Platelet Count:** Patient’s most recent Platelet lab value (10^9^/L) around index diagnosis, up to 6-months pre-index.

**Albumin:** Patient’s most recent albumin (g/dL) lab value around index diagnosis, up to 6-months pre-index.

**Total WBC:** Patient’s most recent total WBC (10^9^/L) lab value around index diagnosis, up to 6-months pre-index.

**Hepatitis:** Hepatitis B/C History. ICD Codes: ICD 10 :B16 (and all sub-codes), B17.1 (and all sub-codes), B17.0, B17.2, B17.8, B17.9, B25.1, B15 (and all sub-codes), B18.0, B18.1, B19.1, B19.11, B18.2, B19.2, B19.20, B19.21, B18.8, B18.9, B19, B19.0, B19.9

ICD 9 : 070.30, 070.20, 070.51, 070.41, 074.8, 573.1, 078.5, 070.59, 070.49, 070.1, 070.0, 070.52, 070.42, 070.53, 070.43, V02.61, 070.32, 070.22, 070.23, 070.33, 070.21, 070.31, V02.62, 070.54, 070.44, 070.54, 070.70, 070.71, 070.9

**SBP:** History of spontaneous bacterial peritonitis. ICD Codes: ICD 10: K65.2 ,ICD 9: 567.23

**Variceal Bleed:**

History of variceal bleed. ICD Codes: ICD 10: I85.01, I85.11, I86.4 , ICD 9: 456.20, 456.0, 456.8

**HE:** Diagnosed history of hepatic encephalopathy. ICD Codes: ICD 9: 572.2, ICD 10: K72.90

**Medications:**

**PPI:** Evidence of Omeprazole, Pantoprazole, Lansoprazole, Esomeprazole, or Rabeprazole prescription, up to 90 days pre-index.

**Statins:** Evidence of Statin (Atorvastatin, Fluvastatin, Lovastatin, Pitavastatin, Pravastatin, Rosuvastatin, Simvastatin) prescription, up to 90-days pre index.

**Lactulose:** Evidence of lactulose prescription up to 90-days pre-index.

**Propranolol:** Evidence of propranolol prescription up to 90-days pre-index.

**Nadolol:** Evidence of nadolol prescription up to 90-days pre-index.

**Carvedilol:** Evidence of carvedilol prescription up to 90-days pre-index.

**Selective Beta-Blocker:** Evidence of Atenolol, Metoprolol, Betaxolol, Bisoprolol, Acebutolol, Nebivolol, or Pindolol prescription, up to 90-days pre index.

**Diuretic:** Evidence of Furosemide, Torsemide, Bumetanide, or spironolactone prescription, up to 90-days pre index.

**Inpatient Variables:**

**Pneumonia on Admission:** Was the patient admitted to the hospital with pneumonia?

ICD Codes: ICD 9: 480-488, 997.31 (and all sub-codes), ICD 10: J13-J18, J95.851 (and all sub-codes)

**Vasopressor Use During Hospital Stay:** Was the patient given norepinephrine, vasopressin, epinephrine, Terlipressin, or Selepressin during their hospital stay?

**HE on admission:** Was the patient admitted to the hospital with hepatic encephalopathy?

ICD Codes: ICD 9: 572.2, ICD 10: K72.90

**Mechanical Ventilation:** Did the patient require mechanical ventilation or other airway support during admission? CPT: 94002 and 94003

**ICU Transfer:** Was the patient transferred to the ICU during their admission?

**Outcomes:**

**All-Cause Mortality:** 30-day all cause mortality. Source for VA-CDW: Vital Status Master / Death Ascertainment File

**Liver Transplant:** 30-day liver transplant rate. ICD Codes: ICD-10 : Z48.23, Z94.4, T86.49, T86.40, T86.43, ICD-9 : V42.7, V58.44

**Liver Transplant or Death:** 30-day all-cause mortality or liver transplant. TriNetX: Composite outcome. VA/CDW: Not assessed due to low transplant rates.

**Transplant-Free Mortality:** 30-day all-Cause mortality, among those who did not receive LVA-CDW: Not assessed due to low transplant rates.

**Supplementary Table 1: Cohort Characteristics**

| **Variable** | **VA-CDW (*n*=2,237)** | | | **TriNetX (*n=*2,291)** | | |
| --- | --- | --- | --- | --- | --- | --- |
|  | **No-Rifaximin**  **(*n* = 2,119, 94.7%)** | **Rifaximin**  **(*n* = 118, 5.3%)** | ***p*-value** | **No-Rifaximin**  **(*n* = 1,648, 71.9%)** | **Rifaximin**  **(*n* = 643, 28.1%)** | ***p*-value** |
| **Demographics** |  |  |  |  |  |  |
| Age (±SD) | 64.29 (± 9.57) | 61.36 (± 8.18) | **<0.001** | 58.89 (± 13.24) | 57.20 (± 11.27) | **0.002** |
| Male Sex | 2043 (97.3%) | 114 (96.6%) | 0.883 | 961 (58.3%) | 351 (54.6%) | 0.116 |
| White Race | 1480 (74.7%) | 77 (72.0%) | 0.608 | 1223 (74.2%) | 451 (70.1%) | 0.057 |
| Hispanic Ethnicity | 180 (8.7%) | 12 (10.6%) | 0.604 | 153 (9.3%) | 65 (10.1%) | 0.599 |
| **Cirrhosis details** |  |  |  |  |  |  |
| Alcohol Etiology | 409 (19.3%) | 21 (17.8%) | 0.777 | 282 (17.1%) | 366 (56.9%) | **<0.001** |
| Chronic Viral Hepatitis Etiology | 1054 (49.7%) | 80 (67.8%) | **<0.001** | 220 (13.4%) | 475 (73.9%) | **<0.001** |
| History of HE | 497 (23.5%) | 104 (88.1%) | **<0.001** | 402 (24.4%) | 260 (40.4%) | **<0.001** |
| History of SBP | 249 (11.8%) | 41 (34.7%) | **<0.001** | 153 (9.3%) | 189 (29.4%) | **<0.001** |
| History of Variceal Bleed | 272 (12.8%) | 49 (41.5%) | **<0.001** | 99 (6.0%) | 177 (27.5%) | **<0.001** |
| Albumin(±SD) | 2.88 (± 0.86) | 2.90 (± 0.88) | 0.787 | 3.09 (± 0.79) | 2.78 (± 0.72) | **<0.001** |
| MELD-Na (±SD) | 19.43 (± 9.29) | 25.06 (± 10.34) | **<0.001** | 15.95 (± 7.50) | 20.20 (± 8.09) | **<0.001** |
| Median WBC median [IQR] | 7.57 [5.14-11.60] | 7.95 [4.93-12.55] | 0.689 | 7.16 [5.09-10.31] | 6.30 [4.10-10.00] | **<0.001** |
| **Medications** |  |  |  |  |  |  |
| PPI | 423 (20.0%) | 59 (50.0%) | **<0.001** | 953 (57.8%) | 551 (85.7%) | **<0.001** |
| Diuretics | 534 (25.2%) | 75 (63.6%) | **<0.001** | 769 (46.7%) | 494 (76.8%) | **<0.001** |
| Statin | 259 (12.2%) | 12 (10.2%) | 0.603 | 498 (30.2%) | 78 (12.1%) | **<0.001** |
| Lactulose | 200 (9.4%) | 71 (60.2%) | **<0.001** | 344 (20.9%) | 591 (91.9%) | **<0.001** |
| Selective Beta-Blocker | 225 (10.6%) | 17 (14.4%) | 0.255 | 538 (32.6%) | 159 (24.7%) | **<0.001** |
| Propranolol | 99 (4.7%) | 22 (18.6%) | **<0.001** | 70 (4.2%) | 67 (10.4%) | **<0.001** |
| Nadolol | 7 (<1.0%) | 2 (1.7%) | 0.126 | 49 (3.0%) | 111 (17.3%) | **<0.001** |
| Carvedilol | 91 (4.3%) | 11 (9.3%) | **0.020** | 230 (14.0%) | 42 (6.5%) | **<0.001** |
| **Inpatient Variables** |  |  |  |  |  |  |
| ICU Transfer | 542 (25.6%) | 44 (37.3%) | **0.007** | 473 (28.7%) | 288 (44.8%) | **<0.001** |
| Mechanical Ventilation | 432 (20.4%) | 23 (19.5%) | 0.906 | 400 (24.3%) | 298 (46.3%) | **<0.001** |
| Shock / Vasopressor Use | 661 (31.2%) | 34 (28.8%) | 0.679 | 213 9 (12.9%) | 180 (28.0%) | **<0.001** |
| HE on Admission | 209 (9.9%) | 23 (19.5%) | **0.001** | 53 (3.2%) | 90 (14.0%) | **<0.001** |
| **30-day Outcomes** |  |  |  |  |  |  |
| All Cause-Mortality | 467 (22.0%) | 44 (37.3%) | **<0.001** | 221 (13.4%) | 201 (31.3%) | **<0.001** |
| Liver Transplant | 55 (2.6%) | 4 (3.4%) | 0.819 | 186 (11.2%) | 156 (24.3%) | **<0.001** |

PPI: proton pump inhibitors, MELD-Na: model for end-stage liver disease-sodium, HE: hepatic encephalopathy, SBP: spontaneous bacterial peritonitis. All data are presented as number (%) unless mentioned otherwise.

**Supplementary Fig. 1A – Liver Transplant Model, TriNetX**

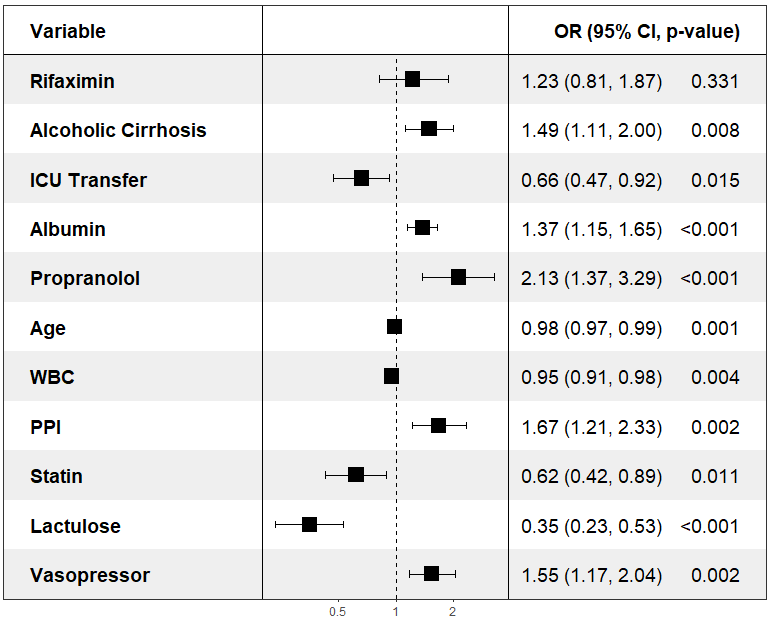
Forest plot with OR and 95% CI for 30-day liver transplant. WBC: white blood cell count, PPI: proton pump inhibitor use, ICU: intensive care unit.

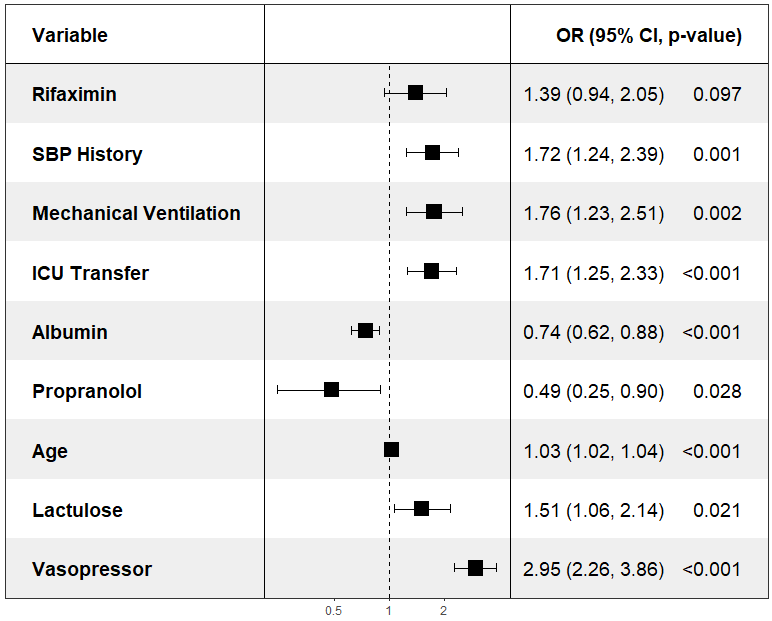
**Supplementary Fig. 1B – 30-Day Death without Transplant, TriNetX**

Forest plot with OR and 95% CI for 30-day death in patients who did not receive a liver transplant. SBP: spontaneous bacterial peritonitis, ICU: intensive care unit.
